# Defining severe acute respiratory infection hospitalisations for national register-based surveillance in Finland, 2022-2025

**DOI:** 10.64898/2026.08.30.26361776

**Authors:** Anthony Ruesta-Maijala, Toni Lehtonen, Jussi Sane, Tuija Leino

## Abstract

**Background:** Severe acute respiratory infections (SARI) strain healthcare systems. Sentinel surveillance remains central to SARI monitoring, but routinely collected hospital discharge data offer a scalable, population-wide complement. In Finland, national registers now enable register-based surveillance, yet SARI case definitions remain unevaluated.

**Aim:** To evaluate whether routinely collected electronic health records can support register-based SARI surveillance and establish a national case definition.

**Methods:** We conducted a retrospective register-based study linking inpatient discharge data from the Finnish Care Register for Health Care (Hilmo) and laboratory-confirmed pathogen notifications from the National Infectious Diseases Register (NIDR). Admissions were aggregated into hospitalisation episodes using generic and pathogen-specific respiratory ICD-10 codes and linked to laboratory-confirmed respiratory pathogens within an admission-centred window. We assessed the impact of diagnostic coding position, laboratory linkage windows and alternative case definitions on age distribution, seasonality and epidemic trend detection.

**Results:** We included 145,435 respiratory hospitalisation episodes. Laboratory confirmations clustered around admission, and a −7-to-+3-day window was selected; 51,498 (35.4%) had a linked laboratory confirmation. Specific primary-position diagnoses preserved clear seasonality and age distributions consistent with SARI epidemiology, whereas secondary-position diagnoses showed attenuated seasonality. A combined case definition incorporating specific primary diagnoses and laboratory-supported syndromic episodes produced stable epidemic curves while improving sensitivity over laboratory confirmation alone.

**Conclusion:** National discharge and laboratory registers can support robust SARI surveillance in Finland when case definitions are carefully designed. A combined register-based definition balances specificity, sensitivity and feasibility, complementing sentinel surveillance and integrated respiratory monitoring.

## INTRODUCTION

Severe acute respiratory infections (SARI), such as those caused by influenza viruses, respiratory syncytial virus (RSV) and coronaviruses, remain a major cause of hospitalisation and mortality worldwide (1, 2). Seasonal epidemics impose a considerable burden on healthcare systems and particularly affect vulnerable populations including children, older adults, and those with underlying health conditions (3–6). To protect public health, robust surveillance systems are required to monitor SARI trends, enable timely detection of epidemic waves and assess the impact of interventions. In Finland, SARI monitoring has traditionally relied on sentinel surveillance. Historically the national hospital discharge records in the Finnish Care Register for Health Care have been updated only once per year, which used to make it unsuitable for non-retrospective surveillance. This has since changed in 2020, when the register was updated to receive records shortly after the discharge from hospital, making data valid for on-line surveillance usage.

For register-based surveillance, SARI case definitions are typically operationalised using hospital discharge data recorded with diagnosis codes such as the International Classification of Diseases (ICD-10), laboratory-confirmed pathogen reports, or record-linked combinations of both. Each approach, however, carries inherent limitations and may introduce potential biases in episode estimates. For example, ICD-coded data offer broad coverage and allow retrospective analyses, but diagnostic codes may vary in specificity and accuracy across hospitals (7–9). In contrast, laboratory-confirmed cases are highly specific but are constrained by testing practices; only a subset of hospitalised patients is tested, and testing policies may fluctuate across seasons and epidemic phases (10–12). In practice, combining ICD-coded diagnoses with laboratory confirmations may improve validity, but requires transparent linkage windows, de-duplication rules, and careful handling of multiple admissions and tests within episodes.

Several studies have evaluated the robustness of register-based SARI surveillance. A five-country study (Denmark, Iceland, Malta, Norway, and Spain) found that ICD-10 codes for COVID-19 and influenza identified SARI cases with good accuracy compared with laboratory results, whereas RSV codes performed poorly, highlighting the need for specific, context-tested definitions (10). In Norway, national register analyses captured distinct epidemic peaks of COVID-19, influenza, and RSV, and emphasised the added value of linking registries with other data sources (13). Similarly, a Danish study showed that ICD-10-based definitions reliably reflected seasonal trends in hospitalisations (14). Despite these advances, there is no consensus on optimal case definitions, as performance remains context-dependent and influenced by coding practices, healthcare utilisation, and testing strategies.

In Finland, no systematic evaluation has yet been conducted to determine how SARI hospitalisations are best identified using national registers. Specifically, it remains unclear how specific versus non-specific ICD-10 codes, alone or in combination with laboratory confirmation, influence disease burden estimates and epidemic trend detection. In this study, by linking data from the Care Register for Health Care (Hilmo) and the Finnish National Infectious Diseases Register (NIDR), we aimed to establish a validated national case definition for SARI that balances accuracy, scalability, EU comparability, and long-term feasibility. We compared candidate case definitions with the goal of identifying an algorithm that supports both timely epidemic detection and accurate estimation of hospitalisation burden.

## METHODS

### Study design and data sources

We conducted a retrospective, register-based observational study using routinely collected national health data from Finland. The study period covered 1 January 2022 to 31 July 2025.

Data on hospitalisations were obtained from the Finnish Care Register for Health Care (Hilmo), which contains nationwide information on inpatient secondary care events, including admission and discharge dates, and diagnoses coded according to the International Classification of Diseases, 10th revision (ICD-10) (15). Laboratory-confirmed infectious disease notifications were obtained from the Finnish National Infectious Diseases Register (NIDR), which records positive laboratory test results for notifiable pathogens defined in the Communicable Disease Act (16).

Data from Hilmo and NIDR were linked at the individual level using unique personal identifiers.

### Study population and episode definition

The study population consisted of secondary care inpatient episodes with at least one respiratory illness ICD-10 diagnosis code recorded in any diagnostic position. Primary health care encounters were not included.

Hospitalisation episodes were constructed by combining consecutive inpatient events for the same individual into a single episode if the gap between discharge and the next admission was ≤ 2 days. Individuals could contribute more than one episode during the study period. All analyses were conducted at the episode level.

### ICD-10 code lists and identification of respiratory illness episodes

Respiratory illness episodes were identified using ICD-10 diagnosis codes recorded at hospital discharge. Two sets of ICD-10 codes were used:

#### Generic respiratory illness codes

A broad set of ICD-10 codes were used to identify episodes for inclusion in the study cohort: J00-J06, J09-J18, J20-J22, J40, J80, J85.1, J86 and J96.0. Episodes with any of these codes recorded in either the primary or secondary diagnostic position were included in the cohort.

#### Specific ICD-10 codes

Used to identify episodes explicitly coded as specific respiratory infections, regardless of laboratory confirmation:

- SARS-CoV-2 (COVID-19): U07.1
- Influenza: J09-J11
- Respiratory syncytial virus (RSV): B97.4, J12.1, J20.5, J21.0
- Adenovirus: B97.0, J12.0

### Laboratory data and pathogen classification

Laboratory-confirmed respiratory pathogen data were obtained from NIDR. The analysis focused on the following pathogens with consistent national reporting: SARS-CoV-2, Influenza A, Influenza B, RSV and Adenovirus.

In addition, laboratory-confirmed detections for 80 other pathogens associated with respiratory symptoms (Supplementary Table 1) were collected and grouped under a single category termed “Other pathogens” reflecting heterogeneous testing practices and coding conventions. No specific ICD-10 code list was defined for these additional pathogens.

### Linkage of laboratory results to hospitalisation episodes

Laboratory-confirmed pathogen results from NIDR were linked to hospitalisation episodes based on the temporal proximity between the date of laboratory confirmation and the episode admission date.

An initial exploratory linkage window of −28 to +28 days relative to the admission date was used to assess the temporal distribution of laboratory confirmations in relation to hospitalisation. Based on this assessment and comparison of alternative candidate windows, a final linkage window of −7 to +3 days relative to the admission date was selected for the main analyses.

An episode was classified as laboratory-confirmed for a given pathogen if any positive laboratory test for that pathogen occurred within the defined linkage window. Multiple laboratory-confirmed pathogens could be linked to the same episode. Episodes with laboratory confirmation for more than one pathogen were therefore allowed to contribute to multiple pathogen-specific analyses.

### Classification of episodes by aetiology

Episodes were categorised based on the presence of laboratory-confirmed pathogens and/or specific ICD-10 codes. An episode could be classified as:

- Laboratory-confirmed for one or more respiratory pathogens
- Specific ICD-10 coding in the absence of laboratory confirmation
- Syndromic respiratory, based on generic respiratory illness ICD-10 codes without specific codes or laboratory confirmation

For pathogen-specific time series analyses, episodes with multiple laboratory-confirmed pathogens were counted separately for each pathogen, resulting in double counting across pathogen-specific curves where applicable.

### Development of candidate SARI case definitions

We specified a set of candidate case definitions to compare alternative approaches for identifying SARI hospitalisations. These comprised seven individual definitions (D1-D7) and two combined definitions (CD1, CD2) formed by uniting complementary definitions.

The candidate definitions ranged from highly specific definitions requiring concordance between specific ICD-10 codes and corresponding laboratory confirmation, to broader syndromic definitions based on respiratory illness ICD-10 codes alone. The combined definitions (CD1 = D2+D3; CD2 = D2+D6) were constructed to balance sensitivity and specificity by incorporating both diagnostic coding and laboratory evidence. Full specifications and the rationale for each definition are presented in Table 1.

**Table 1.**
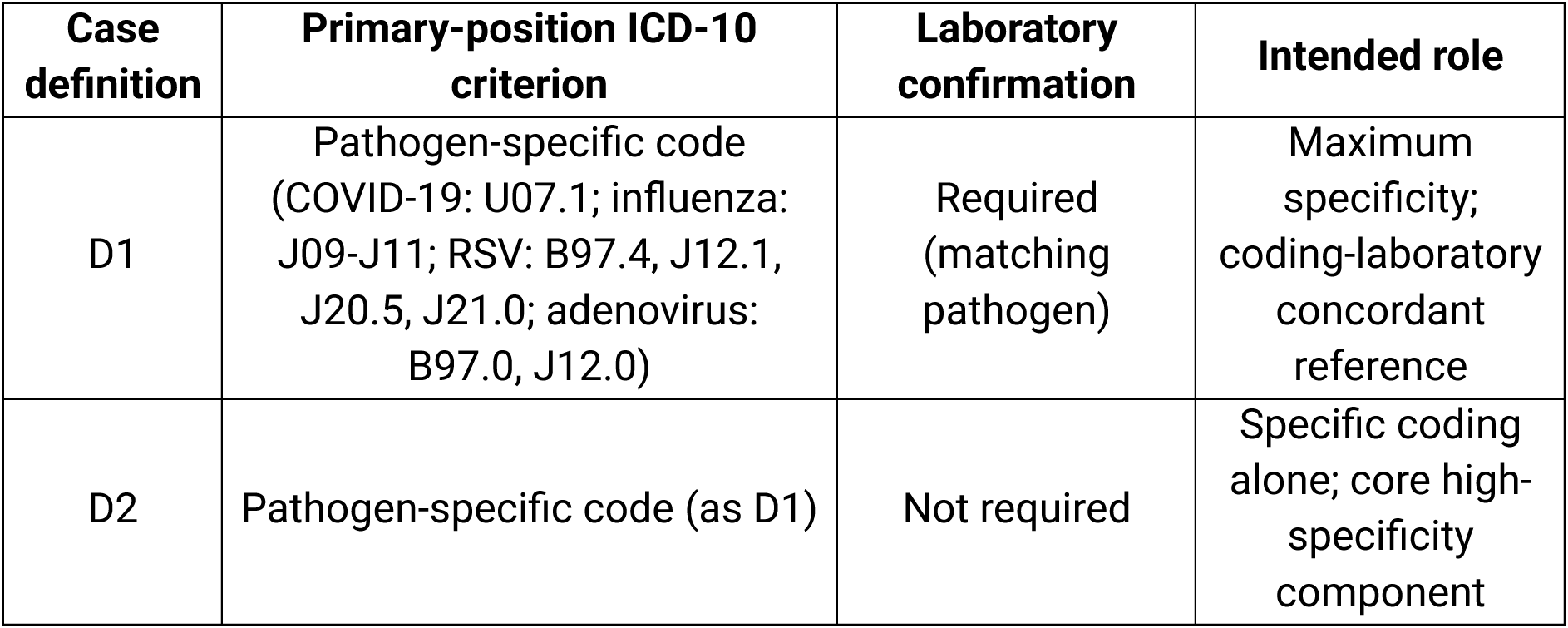
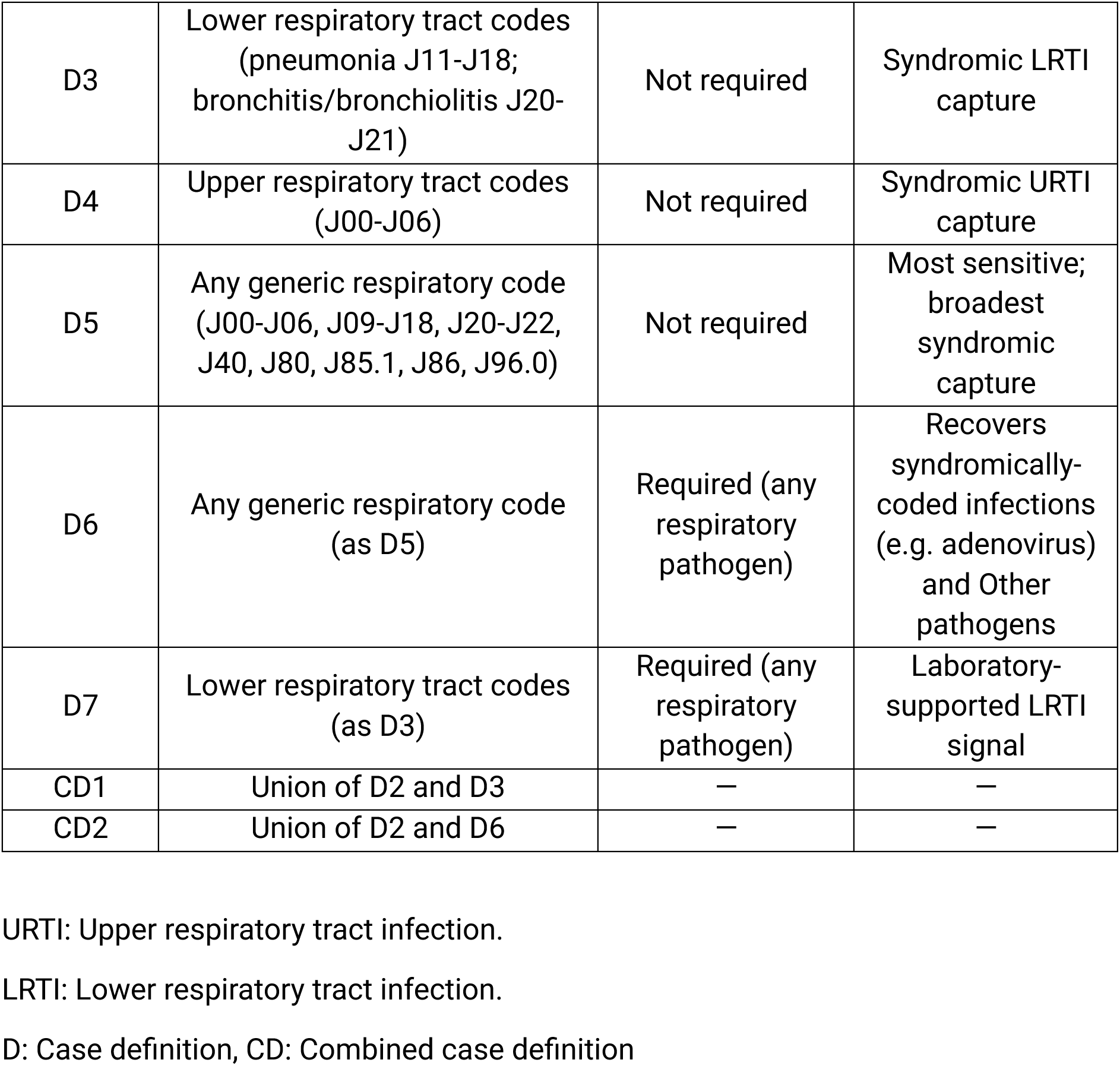
Candidate case definitions evaluated for register-based SARI surveillance.

### Statistical analyses

Descriptive analyses were conducted to characterise the episode-based cohort by age, sex, diagnostic position of ICD-10 codes, laboratory confirmation status, and pathogen group.

Weekly epidemiological time series were constructed for:

- All episodes in the cohort.
- Laboratory-confirmed episodes.
- Episodes with specific ICD-10 coding.
- Episodes captured by each candidate case definition.

Analyses were stratified by age group and diagnostic position (primary vs secondary) where relevant. Comparisons focused on the temporal patterns and seasonal coherence of time series across alternative case definitions.

All analyses were performed using R Statistical Software (v4.5.1; R Core Team 2025).

### Ethics and data protection

This study was conducted using routinely collected national health register data. It has been carried out at the Finnish Institute for Health and Welfare based on the Communicable Diseases Act § 44 (https://www.finlex.fi/api/media/statute-foreign-language-translation/687819/mainPdf/main.pdf?timestamp=2016-12-20T22%3A00%3A00.000Z). Neither specific ethical approval of this study nor informed consent from the participants was needed. Data were pseudonymised prior to analysis, and all analyses were performed in accordance with national data protection regulations.

## RESULTS

This study included 145,435 inpatient hospitalisation episodes recorded in the Care Register for Health Care (Hilmo) between 1 January 2022 and 31 July 2025 that fulfilled the inclusion criteria for respiratory illnesses. Episodes were defined by combining temporally adjacent inpatient events for the same individual into hospitalisation periods, allowing individuals to contribute more than one episode over the study period. All analyses were therefore conducted at the episode level.

### Primary versus secondary position of respiratory illness ICD-10 codes

Episodes were stratified according to whether a respiratory illness ICD-10 diagnosis code was recorded in the primary or secondary diagnostic position at discharge.

Marked differences were observed in the age distribution of episodes between these two groups (Figure 1). Episodes with a diagnosis code recorded in the secondary position increased progressively with age, with the highest number of episodes occurring among older adults. In contrast, episodes with a diagnosis code recorded in the primary position showed a U-shaped age distribution, with higher numbers among young children and older adults.

**Figure 1.**
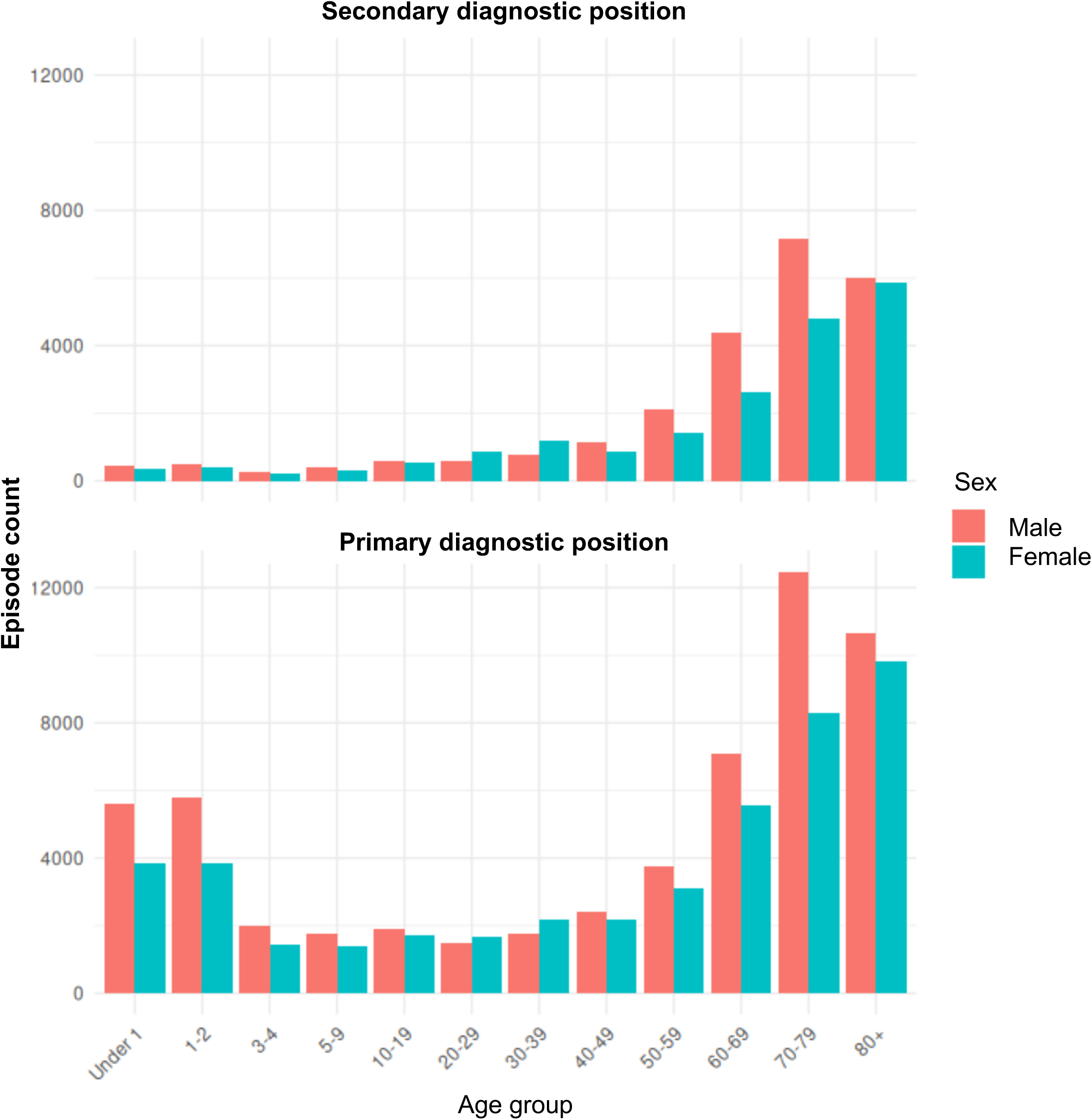
Age and sex distribution of respiratory illness hospitalisation episodes by diagnostic position, Finland, 2022-2025. Age- and sex-specific distribution of hospitalisation episodes with respiratory illness ICD-10 diagnoses recorded in the secondary diagnostic position (upper panel) and primary diagnostic position (lower panel). Bars represent the number of episodes within each age group, stratified by sex.

These differences were consistent across sexes and suggest that the diagnostic position of respiratory illness ICD-10 codes is associated with distinct epidemiological profiles at the population level.

### Timing of laboratory confirmation and selection of the linkage window

To assess the temporal relationship between hospital admission and laboratory testing, the distribution of time from admission to laboratory confirmation was examined among episodes with linked pathogen test results. Analyses were stratified by diagnostic position -primary vs secondary codes-and by pathogen group.

Across both diagnostic-position strata, primary and secondary, laboratory confirmations clustered around the time of admission, with the highest frequency occurring between approximately 4 days before and 7 days after the admission date (Figure 2). This pattern was observed consistently across major respiratory pathogens, including SARS-CoV-2, influenza A and B, RSV, adenovirus, and Other pathogens.

**Figure 2.**
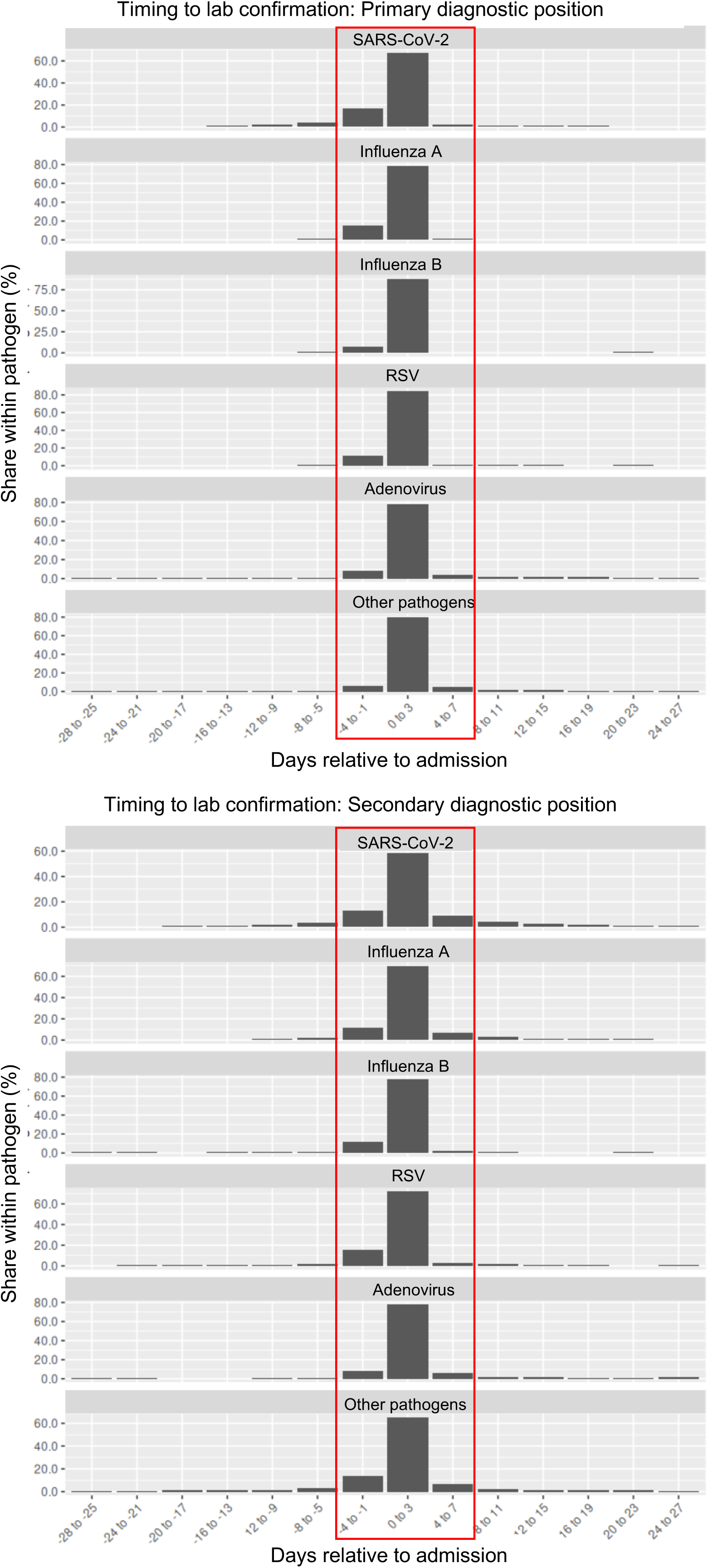
Timing of laboratory confirmation relative to hospital admission, by diagnostic position and pathogen group, Finland, 2022–2025. Distribution of time from hospital admission to laboratory confirmation among episodes with linked pathogen test results, stratified by diagnostic position of respiratory illness ICD-10 codes (primary vs secondary) and by pathogen group. Histograms are shown in 4-day bins, with the y-axis representing the percentage of laboratory-confirmed episodes within each time interval. Across pathogens and diagnostic positions, laboratory confirmations cluster closely around the admission date, with the highest proportions observed between approximately −4 and +7 days relative to admission (red box).

Based on these observations, several candidate admission-to-test linkage windows were compared, including −7 to +3 days, −7 to +7 days, −4 to +7 days, and −3 to +3 days. Weekly epidemiological curves constructed using these alternative windows showed only marginal differences in the number and timing of episodes captured (Figure 3). Analyses of relative changes in episode capture showed that broader windows resulted in only marginal differences in the proportion of episodes compared with the −7 to +3 day window. In line with previous register-based studies in Finland, and given the minimal impact on observed trends, the −7 to +3 day linkage window was retained for all subsequent analyses (Figure 3).

**Figure 3.**
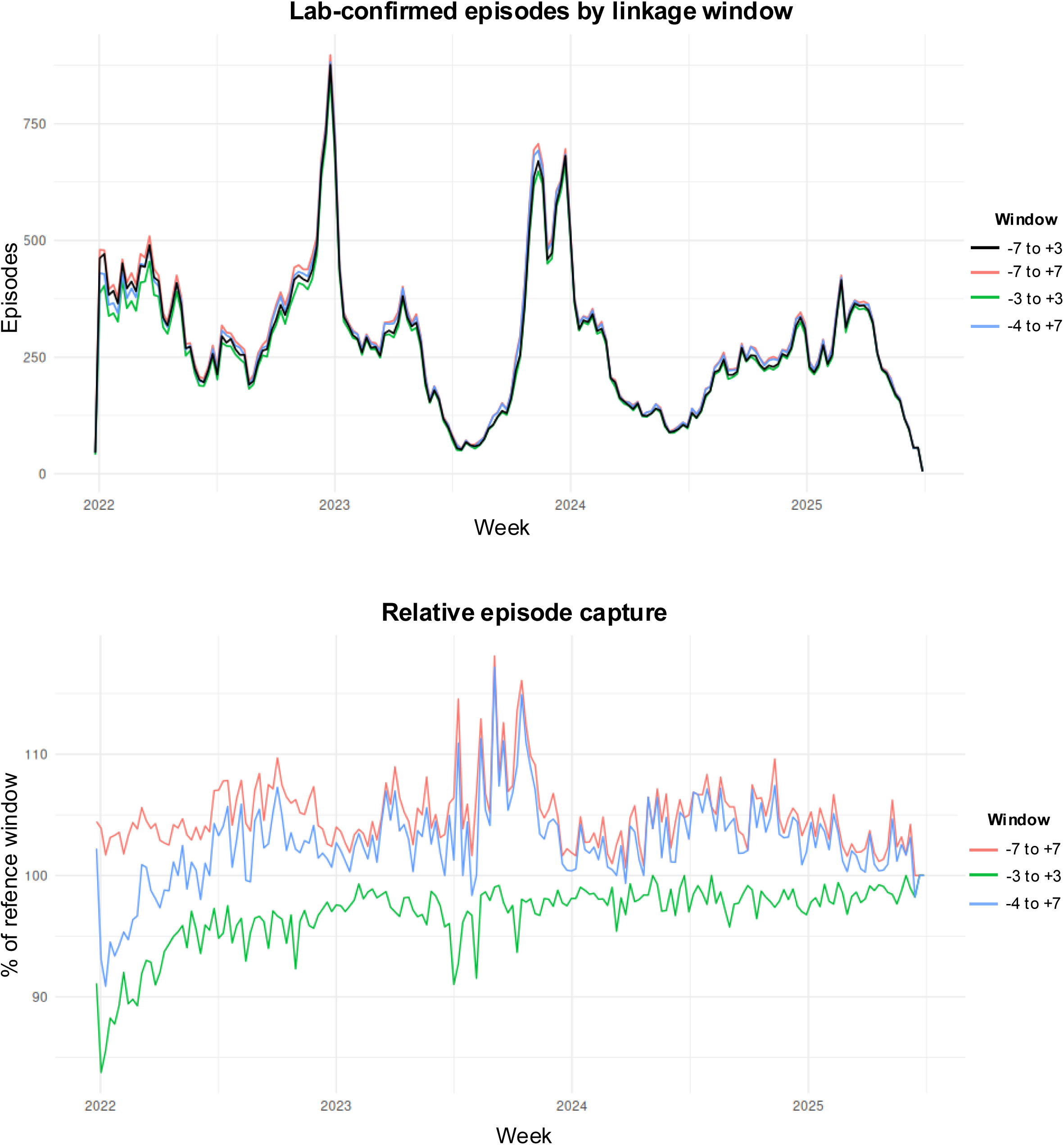
Sensitivity of laboratory-linked episode identification to alternative admission-to-test linkage windows, Finland, 2022–2025. (Upper panel) Weekly time series of episodes with at least one linked laboratory-confirmed respiratory pathogen, constructed using alternative admission-to-test linkage windows (−7 to +3, −7 to +7, −4 to +7 and −3 to +3 days). (Lower panel) Relative episode capture under each alternative linkage window compared with the reference window (−7 to +3 days).

### Laboratory-confirmed pathogen-specific trends as a benchmark

Weekly time series of laboratory-confirmed episodes were constructed for major respiratory pathogens using the selected −7 to +3 day linkage window (Figure 4). In total, 51,498 (35.4%) of all included episodes had at least one laboratory-confirmed pathogen associated with clinical respiratory infections.

**Figure 4.**
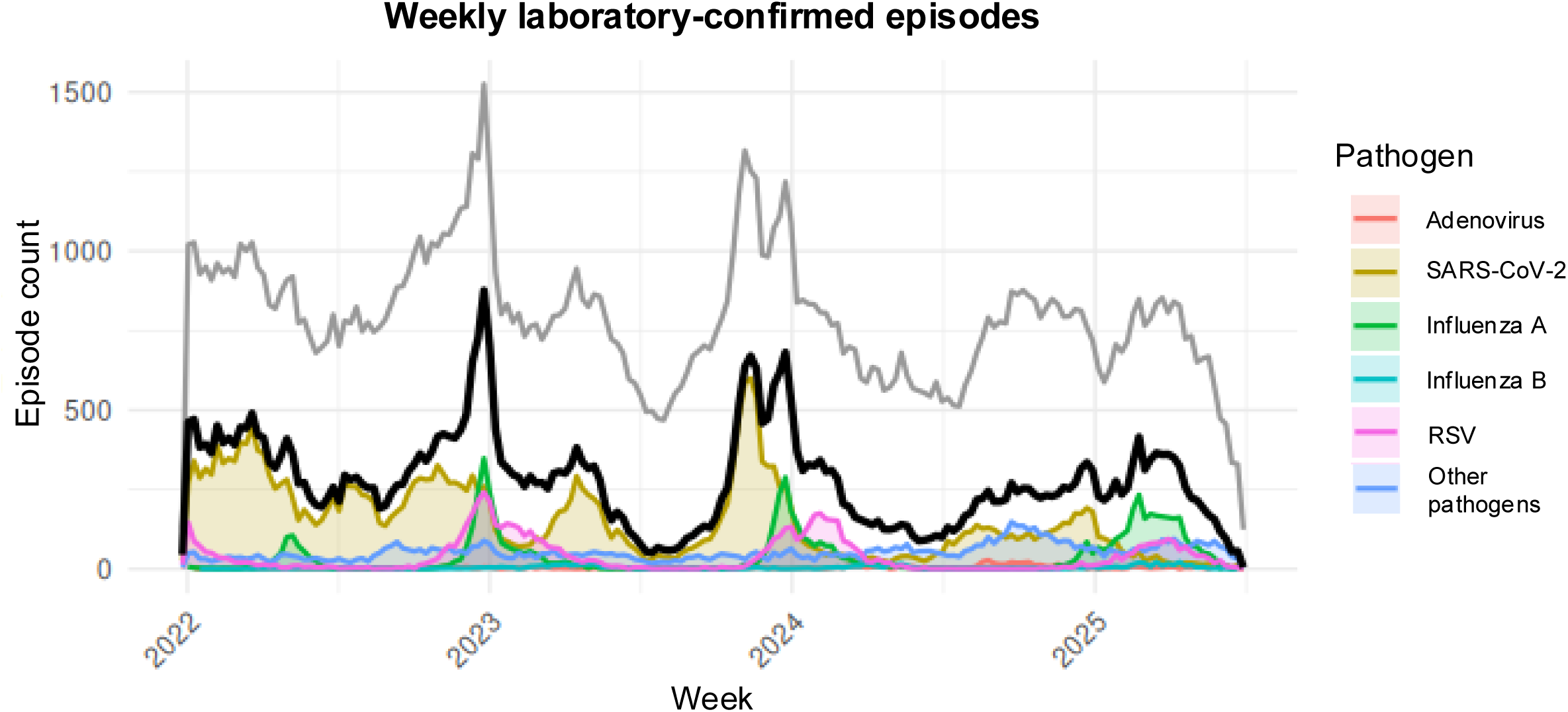
Weekly laboratory-confirmed respiratory episodes as a surveillance benchmark, Finland, 2022-2025. Weekly counts of hospitalisation episodes with laboratory-confirmed respiratory pathogens during the study period (Finland, 2022-2025). The grey line represents all respiratory illness episodes identified from hospital discharge data (study cohort); the black line represents all episodes with any laboratory confirmation; and coloured lines represent pathogen-specific laboratory-confirmed episodes (SARS-CoV-2, influenza A, influenza B, RSV, adenovirus and other known respiratory pathogens).

The combined laboratory-confirmed time series showed clear seasonal variation, with pronounced peaks during winter months across the study period (2022-2025). Age-stratified analyses highlighted heterogeneity in pathogen-specific seasonality, with RSV contributing prominently among younger age groups and influenza and SARS-CoV-2 contributing a larger proportion of episodes among adults and older individuals (Supplementary Figure 1).

To assess the relationship with discharge coding, laboratory-confirmed time series were stratified by whether a corresponding ICD-10 code for the same pathogen was recorded in the primary or secondary diagnostic position (Supplementary Figure 2). Episodes with codes in the primary position preserved seasonal patterns consistent with the overall laboratory-confirmed benchmark, whereas those with codes in the secondary position showed attenuated seasonality.

Given that laboratory confirmation within the linkage window was available for only a subset of episodes (35.4%) in the cohort, laboratory-confirmed time series were used as high-specificity benchmarks of temporal patterns and epidemic dynamics, rather than as estimates of overall SARI hospitalisation burden.

### Concordance between laboratory confirmation and ICD-10 coding

The concordance between laboratory-confirmed respiratory pathogens and corresponding specific ICD-10 codes recorded at hospital discharge was assessed among episodes with and without linked laboratory confirmation (Figure 5). Analyses were restricted to pathogens with established specific ICD-10 codes and consistent coding practices (SARS-CoV-2, influenza A and B, RSV, and adenovirus).

**Figure 5.**
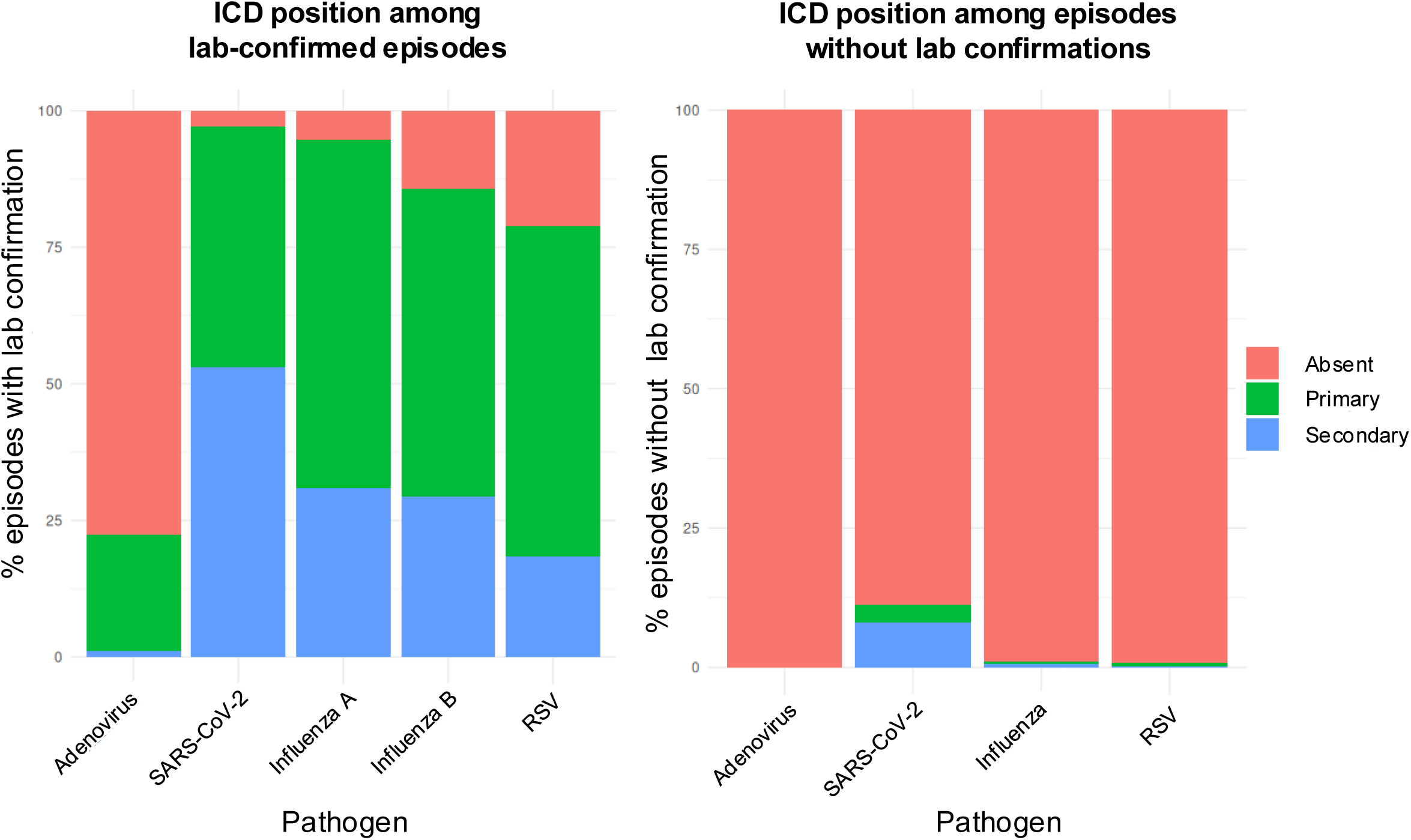
Concordance between laboratory confirmation and pathogen-specific ICD-10 coding, Finland, 2022-2025. Stacked bars show the diagnostic position (primary, secondary, or absent) of pathogen-specific ICD-10 codes recorded at hospital discharge. (Left) Among episodes laboratory-confirmed for each pathogen (SARS-CoV-2, influenza A, influenza B, RSV, adenovirus), the proportion with the corresponding specific code recorded in the primary position, the secondary position, or absent. (Right) Among all episodes without a linked laboratory confirmation, the proportion carrying each pathogen-specific ICD-10 code (SARS-CoV-2, influenza, RSV, adenovirus) in the primary or secondary position, or absent.

Among episodes with laboratory-confirmed SARS-CoV-2, influenza A, influenza B, and RSV infection, more than 75% had a matching specific ICD-10 code recorded in either the primary or secondary diagnostic position. For SARS-CoV-2 laboratory-confirmed episodes, approximately 50% had a matching ICD-10 code recorded as the primary diagnosis. In contrast, concordance was substantially lower for adenovirus, with only ∼25% of laboratory-confirmed adenovirus episodes assigned an adenovirus-specific ICD-10 code; the majority were coded using non-specific respiratory diagnoses (Figure 5).

Among episodes without linked laboratory confirmation, specific ICD-10 codes were rarely recorded for influenza, RSV and adenovirus. For SARS-CoV-2, however, a small proportion (∼10%) of episodes without linked laboratory confirmation carried COVID-19-specific ICD-10 codes.

### Primary diagnoses among laboratory-confirmed episodes with secondary-position specific codes

To evaluate whether secondary-position specific ICD-10 codes reflect respiratory-related admissions, we examined the primary diagnoses of laboratory-confirmed episodes in which the corresponding code appeared only as a secondary diagnosis.

Among laboratory-confirmed SARS-CoV-2 and influenza episodes with specific codes recorded in the secondary position, more than half had a non-respiratory primary diagnosis, suggesting that these admissions were often not primarily due to respiratory illnesses. In contrast, most adenovirus episodes had a respiratory (J-category) primary diagnosis, consistent with syndromic respiratory coding. For RSV, approximately 60% of episodes had a respiratory primary diagnosis (Supplementary Figure 3).

In absolute numbers, laboratory-confirmed SARS-CoV-2 episodes substantially outnumbered those for influenza and RSV. Including secondary-position specific ICD-10 codes would therefore disproportionately increase the contribution of SARS-CoV-2 and introduce heterogeneity in primary admission diagnoses, without improving the epidemiological signal. These findings suggest limited added value of secondary-position codes for routine register-based SARI surveillance.

### Comparison of candidate SARI case definitions and final selection

A set of ICD-10- and laboratory-based SARI case definitions was evaluated to identify a definition suitable for routine register-based surveillance in Finland (Table 1).

When examined as weekly time series, several candidate definitions showed limited or inconsistent seasonal variation. In particular, episodes defined solely by upper respiratory tract infection (URTI) codes (D4) exhibited low episode counts and lacked clear seasonal patterns. Definitions based on broad non-specific respiratory ICD-10 codes, J-category (D5, D6) or lower respiratory tract infection (LRTI) (D3, D7), captured substantially larger numbers of episodes but closely approximated the total cohort, with reduced discrimination of epidemic peaks (Figure 6A).

**Figure 6.**
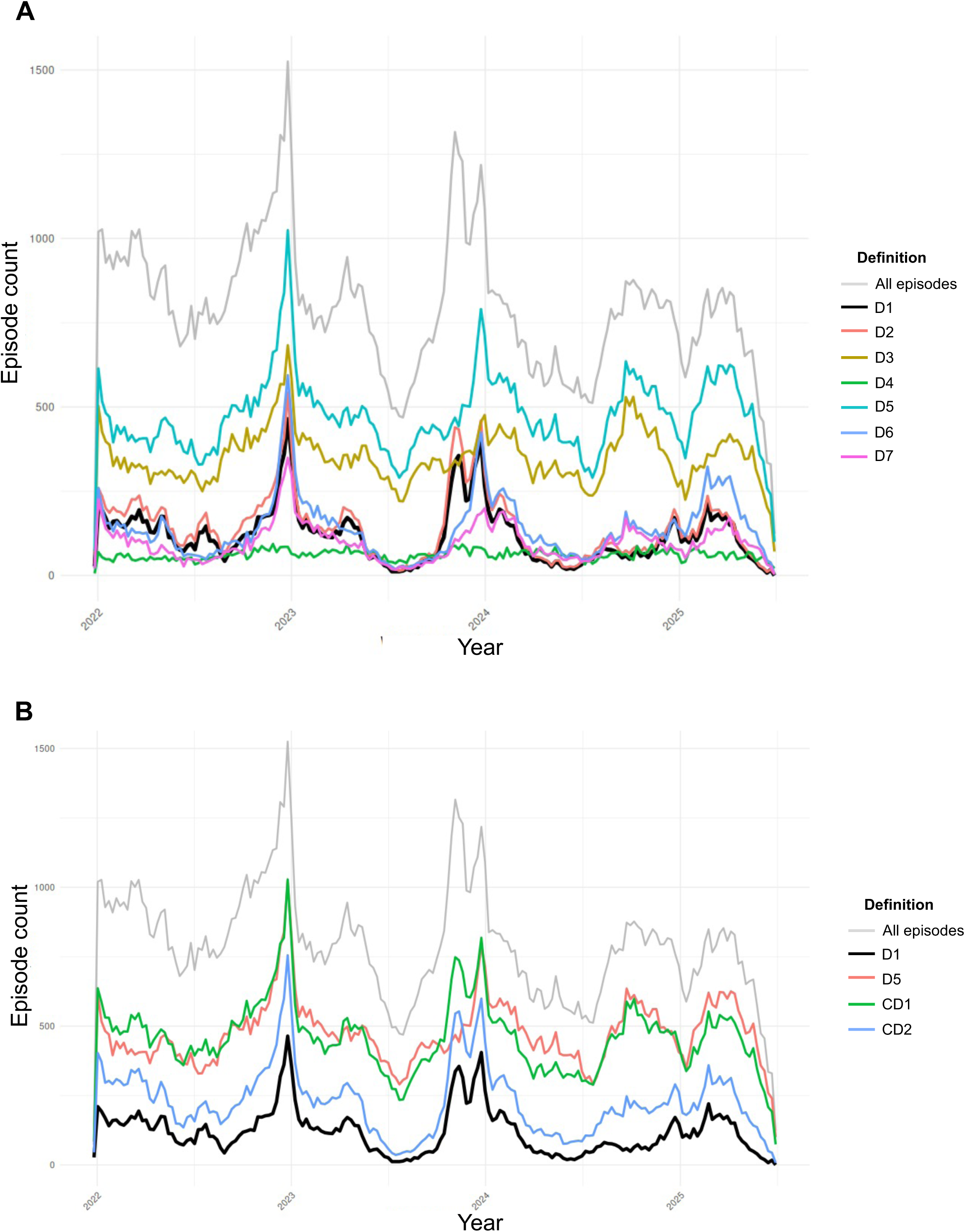
Comparison of candidate SARI case definitions, Finland, 2022-2025. Weekly time series comparing alternative ICD-10- and laboratory-based candidate SARI case definitions. (A) Weekly curves for the seven individual candidate definitions (D1-D7) and the total respiratory illness cohort (grey). (B) The two combined definitions -CD1 (D2+D3) and CD2 (D2+D6)-compared against the high-specificity D1 definition (black) and the total respiratory illness cohort (grey).

Definitions incorporating specific primary ICD-10 codes (D1, D2) demonstrated clearer seasonal patterns aligned with the laboratory-confirmed benchmark but captured a smaller proportion of total episodes. Expanding these definitions by including episodes with respiratory illness ICD-10 codes (J-category) and a timely laboratory-confirmed respiratory pathogen (D6) increased the number of captured episodes while preserving seasonal signal coherence (CD2) (Figure 6B).

Across age groups, CD2 showed consistent temporal patterns that remained distinct from the total number of episodes in our cohort (Supplementary Figure 4). Based on these comparisons, CD2 was selected for further use as a pragmatic, scalable case definition for register-based SARI surveillance.

## DISCUSSION

In this study, we demonstrate the feasibility of a register-based national SARI surveillance strategy in Finland, linking inpatient discharge diagnoses from the Finnish Care Register for Health Care (Hilmo) with laboratory notifications from the National Infectious Diseases Register (NIDR). This enables surveillance using routinely collected data while allowing individuals to contribute multiple hospitalisation episodes over time, reflecting real-world recurrent admissions and reinfections. Similar episode-construction approaches have been used in other Nordic register-based SARI systems to avoid double counting when records are generated at multiple contact points during a single stay (14). Beyond feasibility, a standardised register-based definition would enable consistent monitoring of seasonality and age-specific burden and support applications such as nowcasting, intervention evaluation and cost-effectiveness analyses. Notably, during the COVID-19 pandemic, hospitalisation-based metrics proved more stable than case-based metrics, which are sensitive to changes in testing and healthcare-seeking behaviour (17–20).

Within this framework, the diagnostic position of respiratory ICD-10 codes materially influenced the epidemiological signal. Episodes with codes in the primary position showed pronounced seasonality and an age distribution consistent with established SARI epidemiology (21), whereas secondary-position episodes lost seasonality and skewed older. This is consistent with prior register-based studies showing that including secondary diagnoses increases sensitivity but attenuates seasonality, reducing specificity for epidemic wave detection (22). Secondary positions likely capture more heterogeneous contexts, particularly older patients with multimorbidity in whom respiratory infection precipitates admission through exacerbation of a chronic condition and is coded secondarily (23). Restricting surveillance indicators to primary positions may therefore be more appropriate for routine wave monitoring and directly attributable hospital burden, while secondary diagnoses may still matter for broader burden and healthcare-utilisation assessments, where respiratory infection contributes to admission without being the primary reason.

Our concordance analyses showed that for high-impact respiratory viruses, laboratory-confirmed infections were often reflected in specific ICD-10 discharge coding, particularly for SARS-CoV-2 and influenza and, to a lesser extent, RSV. This suggests that Finnish discharge coding reliably captures clinically relevant infections and supports pathogen-specific primary codes as a high-specificity surveillance component. Adenovirus, by contrast, showed substantially lower concordance (∼25%), consistent with evidence that virus-specific ICD coding is only moderately sensitive for this pathogen (24), likely reflecting clinical salience at discharge, co-detection, and syndromic coding when adenovirus is not the principal cause of admission (25). Adenovirus-attributable burden is therefore better captured by syndromic respiratory definitions combined with laboratory confirmation than by pathogen-specific coding alone.

Integrating these findings, we selected a pragmatic case definition combining (i) specific primary ICD-10 codes and (ii) syndromic respiratory episodes supported by timely laboratory confirmation (including Other pathogens). This balances feasibility and scalability with improved signal-to-noise, recognising that laboratory confirmation alone captured only a minority of respiratory-related episodes (∼35%) and would substantially underestimate counts if used in isolation.

Our findings align with a growing European evidence base showing that electronic health records and administrative registers can support routine SARI surveillance, though optimal proxy definitions depend on national coding and testing practices. In Denmark, a national register-based system using linked registry and microbiology data showed that ICD-10-based definitions reliably capture weekly trends with short reporting delays, while broader code sets including URTI diagnoses increased sensitivity but reduced specificity and signal clarity, leading to their exclusion from the final definition (14). Consistent with this, URTI codes in our data contributed little to case capture and did not improve seasonal coherence. Norway, in contrast, implemented a more inclusive definition incorporating URTI and LRTI codes across both diagnostic positions, prioritising burden estimation over specificity (13); this achieved higher laboratory coverage (∼52% vs ∼35% in Finland) but introduced greater heterogeneity and required nowcasting and alternative definitions to improve timeliness. Given our objective of early epidemic detection and stable week-to-week monitoring, we prioritised a more constrained definition that reduces heterogeneity over maximising inclusive burden capture.

Consistent with a five-country validation study, ICD-10 codes for COVID-19 and influenza performed well against laboratory-confirmed SARI, whereas RSV and other pathogens were captured less consistently (10), supporting the use of specific primary codes as high-specificity identifiers for SARS-CoV-2 and influenza while supplementing poorer-coded pathogens, notably adenovirus, with laboratory-linked syndromic episodes.

SARI surveillance implementation remains heterogeneous across Europe. An evaluation of 27 countries in 2022-2023 found diverse strategies and data sources, underlining the need for transparent national definitions and continued work towards comparability (26). Our findings reinforce the importance of explicitly evaluating definitional choices (diagnostic position, linkage windows and case definition) when building fit-for-purpose register-based systems.

Some methodological considerations apply. Fixed, admission-anchored linkage windows cannot fully distinguish community-acquired from healthcare-acquired infections, particularly for longer stays or delayed testing. As ECDC guidance notes, SARI surveillance focuses on severe outcomes rather than attribution of infection acquisition, and healthcare-associated infections may be included when severe enough to warrant hospitalisation (27); our approach therefore prioritises temporal proximity and clinical relevance over infection origin.

Further limitations relate to misclassification and incomplete linkage. Wider windows may attribute hospitalisations to preceding viral infections that instead reflect downstream complications, such as bacterial infection following influenza or COVID-19 (28–31), whereas narrower windows may under-link infections tested later in admission or subject to reporting delays. Increasing use of point-of-care tests, which may inform coding but are not captured in laboratory registers, may also lead to under-ascertainment of laboratory-confirmed episodes.

Our study shows how routine national registers can support sustainable, scalable SARI monitoring alongside sentinel surveillance. A case definition is operationally feasible, relying on routinely available ICD-10 diagnoses and laboratory notifications, and allows flexible use: broader definitions for weekly situational awareness, more specific ones for pathogen-focused analyses.

The Finnish definition is a transparent proxy compatible with ECDC guidance, which recognises broader or electronic-record-based SARI definitions when clearly specified (27), and aligns with the European transition towards integrated multi-pathogen surveillance, exemplified by the European Respiratory Virus Surveillance Summary (ERVISS) (32, 33). The included “Other pathogens” laboratory category further provides a pragmatic mechanism to capture severe respiratory episodes beyond the traditionally prioritised viruses, acknowledging variability in coding and testing.

Several steps could strengthen the system without altering its core design. First, timeliness and completeness should be formally evaluated using delayed data extracts to determine an evidence-based reporting lag. Second, routine monitoring for coding-practice drift and laboratory-ICD concordance is recommended. Third, pathogen-specific refinements are warranted where coding performance varies, particularly for RSV and adenovirus. Where feasible, future extensions could incorporate intensive care and mortality outcomes to enhance severity assessment, provided trend stability and timeliness are maintained.

## Data Availability

All data produced in the present study are available upon reasonable request to the authors.

