## Supplementary material for "Defining severe acute respiratory infection hospitalisations for national register-based surveillance in Finland, 2022-2025"

**Supplementary Table 1. Laboratory-confirmed pathogens grouped under the "Other pathogens" category, Finnish National Infectious Diseases Register.**

| Abbreviation | |
|---|---|
| Betahemolyyttinen streptokokki | Legionella pneumophila ssp. Fraseri |
| Bokavirus | Legionella pneumophila ssp. pascullei |
| Bordetella parapertussis | Legionella pneumophila ssp. ssp. pneumophila |
| Bordetella pertussis | Legionella sp. |
| Bordetella sp. | Metapneumovirus |
| Bordetella-like sp. | Moraxella catarrhalis |
| Brucella abortus | Moraxella sp. |
| Brucella canis | Mycobacterium tuberculosis |
| Brucella melitensis | Mycobacterium tuberculosis complex |
| Brucella sp. | Mycoplasma pneumoniae |
| Brucella suis | Mycoplasma sp. |
| Chlamydia pneumoniae | Parainfluenssavirus |
| Chlamydia psittaci | Parainfluenssavirus 4 |
| Chlamydia sp. | Parvovirus |
| Coronavirus | Pneumocystis carinii |
| Corynebacterium diphtheriae | Rinovirus |
| Coxiella burnetiid | RS-virus |
| Coxsackie A virus | SARS-virus |
| Coxsackie B virus | Staphylococcus aureus |
| Echinococcus granulosus | Staphylococcus lajeja, koagulaasi-negatiivisia |
| Echinococcus multilocularis | Staphylococcus laji, koagulaasi-negatiivinen |
| Echinococcus sp. | Streptococcus infantis |
| Echovirus | Streptococcus pneumoniae |
| Ehrlichia sp. | Streptococcus pseudopneumoniae |
| Eikenella corrodens | Streptokokki ryhmä B |
| Enterovirus | Streptokokki ryhmä C |
| Francisella tularensis | Streptokokki ryhmä D |
| Haemophilus influenzae | Streptokokki ryhmä F |
| Haemophilus parahaemolyticus | Streptokokki ryhmä G |
| Haemophilus parainfluenzae | Streptococcus pyogenes |
| Haemophilus paraphrophilus | Legionella longbeachae |
| Haemophilus sp. | Legionella anisa |
| Influenssa A -virus | Pseudomonas aeruginosa |
| Influenssa B -virus | Streptococcus dysgalactiae ssp. equisimilis |
| Influenssavirus | Streptococcus suis |
| Klebsiella pneumoniae | Streptococcus equi ssp. equi |

| | |
|---|---|
| Klebsiella pneumoniae ssp. ozaenae | Streptococcus equi ssp. zooepidemicus |
| Klebsiella sp. | |
| Kokkivaltaista mikrobistoa | |
| Koronavirus 229E | |
| Koronavirus NL63 | |
| Koronavirus OC43 | |
| Legionella pneumophila | |

### Supplementary Figure 1. Age-stratified weekly laboratory-confirmed respiratory episodes by pathogen, Finland, 2022-2025

Weekly time series of laboratory-confirmed respiratory hospitalisation episodes, stratified by age group (<1, 1-2, 3-4, 5-9, 10-19, 20-29, 30-39, 40-49, 50-59, 60-69, 70-79 and ≥80 years). Within each panel, the grey line represents all respiratory illness episodes identified from hospital discharge data (study cohort); the black line represents episodes with any laboratory confirmation; and coloured lines represent pathogen-specific laboratory-confirmed episodes (SARS-CoV-2, influenza A, influenza B, RSV, adenovirus and Other pathogens).

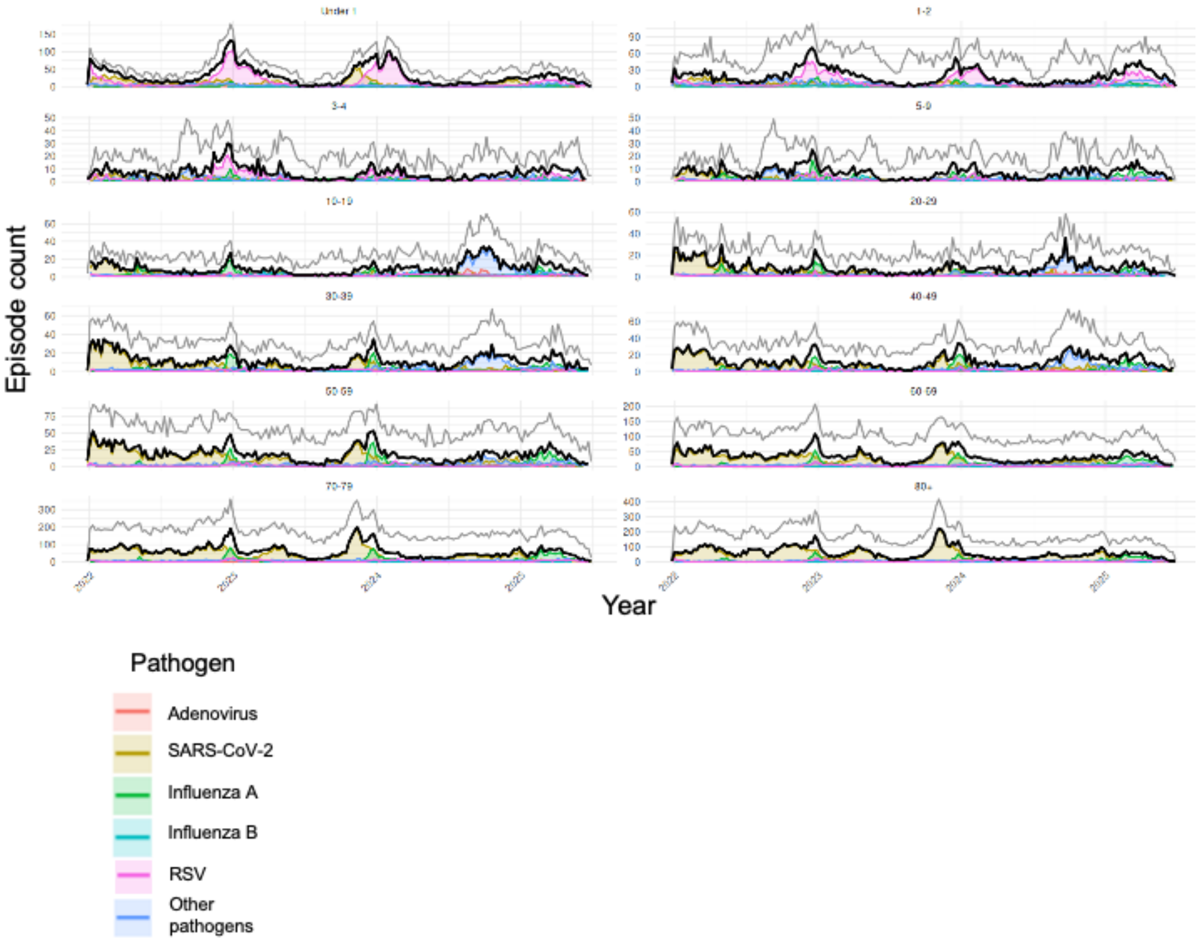

### Supplementary Figure 2. Seasonality of laboratory-confirmed respiratory episodes by diagnostic coding position, Finland, 2022-2025

Weekly time series of laboratory-confirmed respiratory episodes that carried a corresponding pathogen-specific ICD-10 code, stratified by the diagnostic position of that code. Upper panel: code recorded in the primary diagnostic position. Lower panel: code recorded in the secondary diagnostic position.

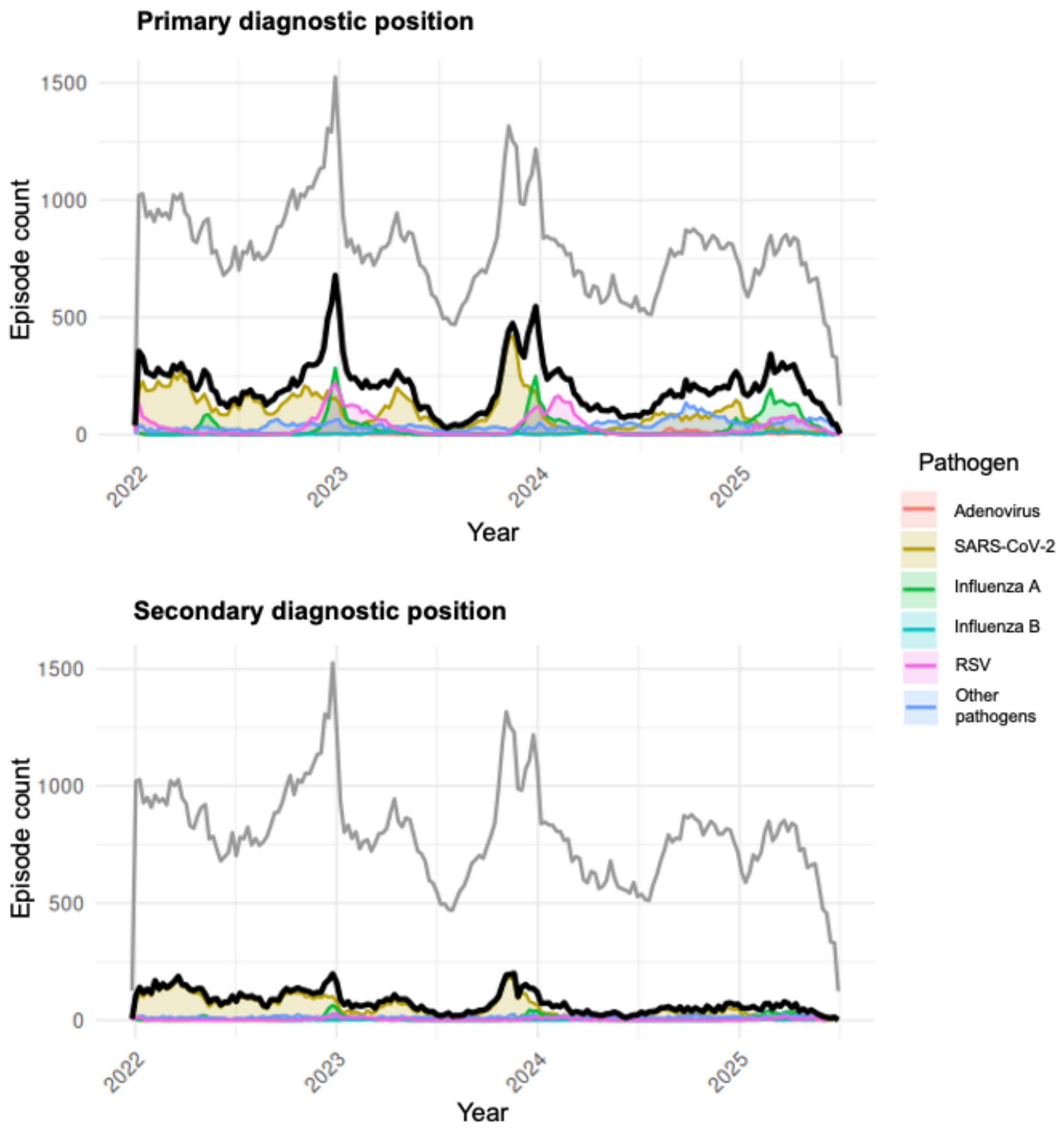


### Supplementary Figure 3. Primary diagnosis categories among laboratory-confirmed episodes with pathogen-specific ICD-10 codes in the secondary position, Finland, 2022-2025

Distribution of primary diagnosis categories among laboratory-confirmed episodes in which the pathogen-specific ICD-10 code was recorded in the secondary diagnostic position, for selected pathogens (SARS-CoV-2, influenza, RSV and adenovirus). For each pathogen, the left bar shows the proportion of episodes with a respiratory (J-category) primary diagnosis (pink) versus a non-respiratory primary diagnosis (blue); the right bar shows the corresponding absolute number of episodes.

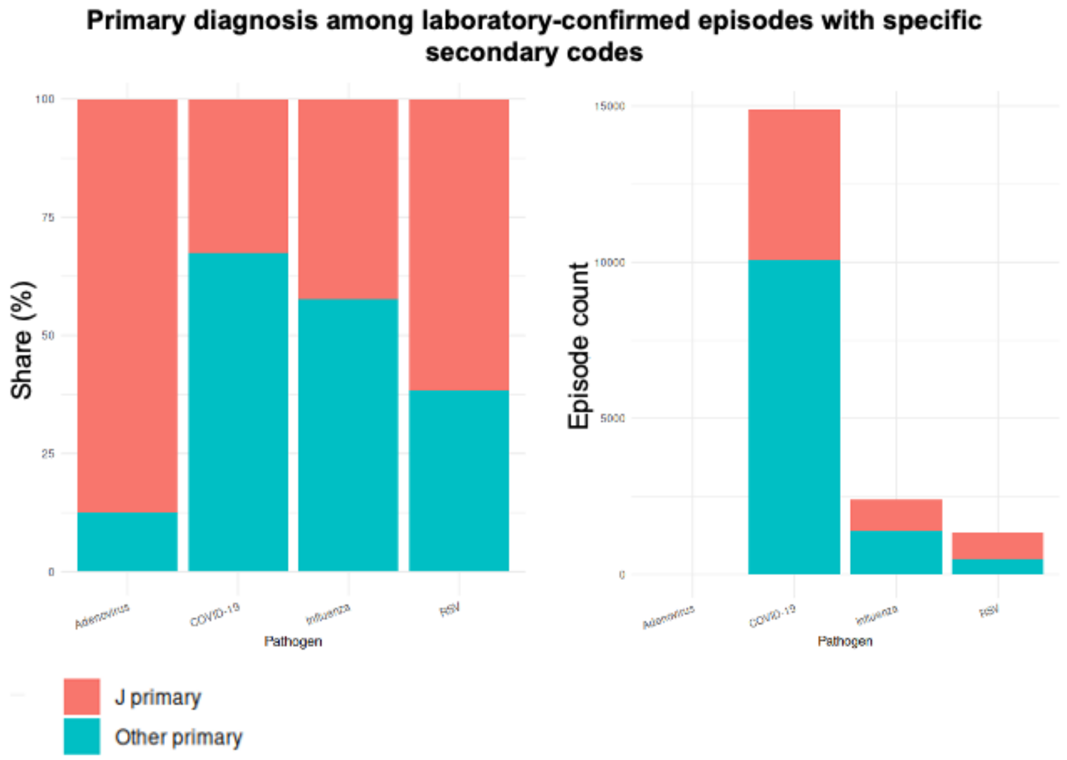

### Supplementary Figure 4. Performance of candidate SARI case definitions by age group, Finland, 2022-2025

Weekly time series for the most sensitive individual definition (D5) and the two combined definitions -CD1 (D2+D3) and CD2 (D2+D6)- stratified by age group (<1, 1-2, 3-4, 5-9, 10-19, 20-29, 30-39, 40-49, 50-59, 60-69, 70-79 and ≥80 years). Within each panel, coloured curves show D5, CD1 and CD2; the grey curve represents the total respiratory illness episodes in that age group (study cohort) and the black curve the episodes captured by the high-specificity D1 definition.

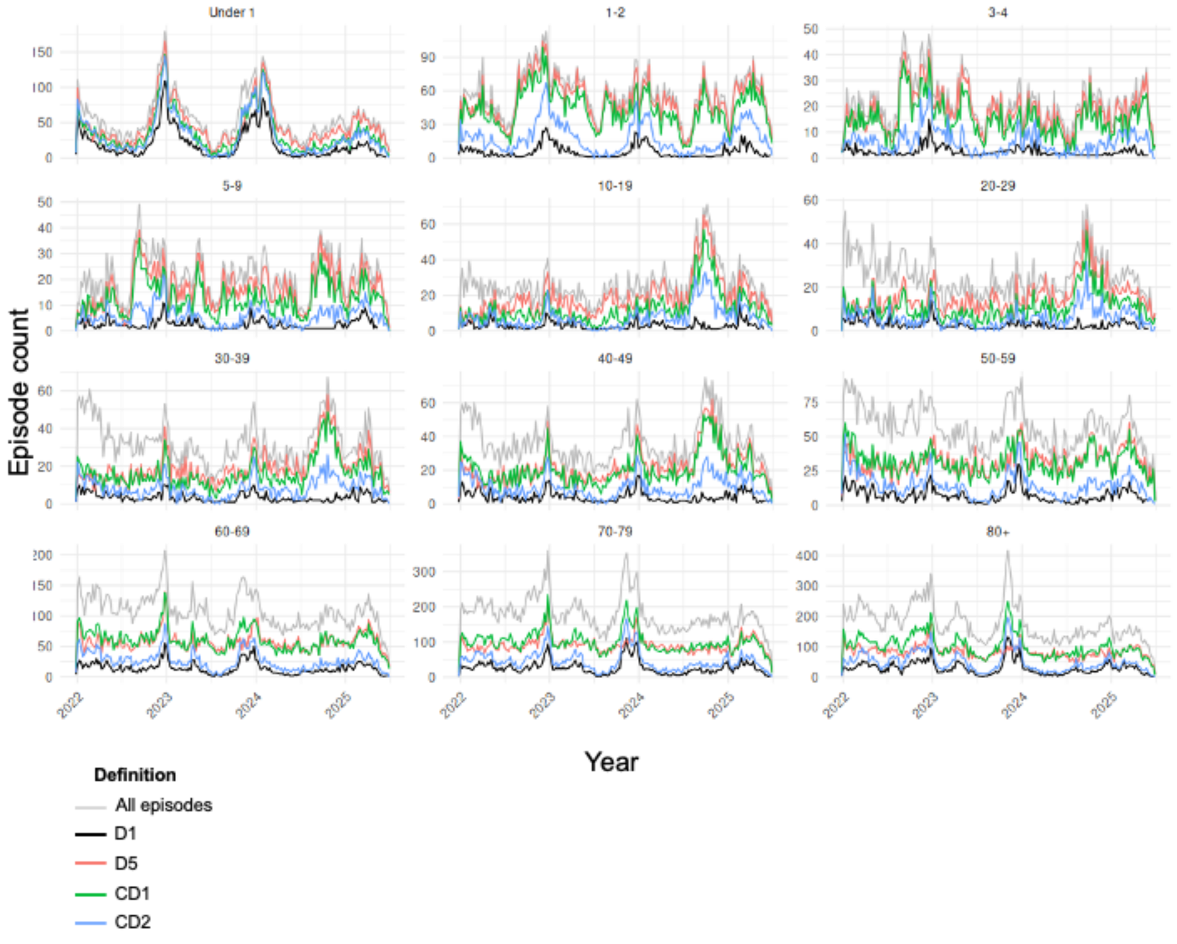
